# A Prospective Study of Long-Term Outcomes Among Hospitalized COVID-19 Patients with and without Neurological Complications

**DOI:** 10.1101/2021.03.18.21253881

**Authors:** Jennifer A. Frontera, Dixon Yang, Ariane Lewis, Palak Patel, Chaitanya Medicherla, Vito Arena, Taolin Fang, Andres Andino, Thomas Snyder, Maya Madhavan, Daniel Gratch, Benjamin Fuchs, Alexa Dessy, Melanie Canizares, Ruben Jauregui, Betsy Thomas, Kristie Bauman, Anlys Olivera, Dhristie Bhagat, Michael Sonson, George Park, Rebecca Stainman, Brian Sunwoo, Daniel Talmasov, Michael Tamimi, Yingrong Zhu, Jonathan Rosenthal, Levi Dygert, Milan Ristic, Haruki Ishii, Eduard Valdes, Mirza Omari, Lindsey Gurin, Joshua Huang, Barry M. Czeisler, D. Ethan Kahn, Ting Zhou, Jessica Lin, Aaron S. Lord, Kara Melmed, Sharon Meropol, Andrea B. Troxel, Eva Petkova, Thomas Wisniewski, Laura Balcer, Chris Morrison, Shadi Yaghi, Steven Galetta

## Abstract

**Background:** Little is known regarding long-term outcomes of patients hospitalized with COVID-19.

**Methods:** We conducted a prospective study of 6-month outcomes of hospitalized COVID-19 patients. Patients with new neurological complications during hospitalization who survived were propensity score-matched to COVID-19 survivors without neurological complications hospitalized during the same period. The primary 6-month outcome was multivariable ordinal analysis of the modified Rankin Scale(mRS) comparing patients with or without neurological complications. Secondary outcomes included: activities of daily living (ADLs;Barthel Index), telephone Montreal Cognitive Assessment and Neuro-QoL batteries for anxiety, depression, fatigue and sleep.

**Results:** Of 606 COVID-19 patients with neurological complications, 395 survived hospitalization and were matched to 395 controls; N=196 neurological patients and N=186 controls completed follow-up. Overall, 346/382 (91%) patients had at least one abnormal outcome: 56% had limited ADLs, 50% impaired cognition, 47% could not return to work and 62% scored worse than average on ≥1 Neuro-QoL scale (worse anxiety 46%, sleep 38%, fatigue 36%, and depression 25%). In multivariable analysis, patients with neurological complications had worse 6-month mRS (median 4 vs. 3 among controls, adjusted OR 2.03, 95%CI 1.22-3.40, P=0.01), worse ADLs (aOR 0.38, 95%CI 0.29-0.74, P=0.01) and were less likely to return to work than controls (41% versus 64%, P=0.04). Cognitive and Neuro-QOL metrics were similar between groups.

**Conclusions:** Abnormalities in functional outcomes, ADLs, anxiety, depression and sleep occurred in over 90% of patients 6-months after hospitalization for COVID-19. In multivariable analysis, patients with neurological complications during index hospitalization had significantly worse 6-month functional outcomes than those without.

## INTRODUCTION

Acute neurological complications of hospitalized COVID-19 patients have been described in several cohorts worldwide^1-9^, however, limited data exists regarding long-term cognitive and functional outcomes. Reports of prolonged memory disorders, fatigue, and persistent respiratory symptoms have been grouped together in the rubric of “long COVID” or “long-hauler” syndrome, yet little is known regarding the prevalence, risk factors or pathophysiology behind this group of symptoms.

Recently, we reported on a prospective cohort of 4,491 COVID-19 patients hospitalized during the spring 2020 surge in New York City and identified 606 (14%) with new neurological disorders during hospitalization^10^. In this study, we followed this initial cohort longitudinally for 6-months to evaluate longer-term outcomes. Our primary aim was to compare global functional outcomes between COVID-19 hospital survivors with and without neurological complications using an ordinal analysis of the modified Rankins Scale (mRS). Secondary outcomes included assessments of activities of daily living, return to work, cognitive function, anxiety, depression, fatigue and sleep abnormalities. We hypothesized that long-term functional outcomes would be worse among patients with neurological complications compared to age, gender and severity of illness-matched COVID-19 controls without neurological complications.

## METHODS

### Study Design and Patient Cohort

We conducted a prospective, observational study of consecutive COVID-19 patients hospitalized at four New York City area hospitals within the same hospital system between March 10, 2020 and May 20, 2020. Inclusion criteria were: age ≥18 years, hospital admission, reverse-transcriptase-polymerase-chain-reaction (RT-PCR) positive SARS-CoV-2 infection from nasopharyngeal sampling, survival to discharge and consent to participate in a follow-up interview. Exclusion criteria were: negative or missing SARS-CoV-2 RT-PCR test, or evaluation in an outpatient or emergency department setting only. Only index admissions were included; readmissions were excluded to avoid double counting. Patients were prospectively screened following hospital admission according to previously published criteria^10^. Briefly, initial screening for inclusion was performed by the emergency department or admitting team, wherein a neurology consult would be triggered according to routine protocol for patients with new or worsened neurological disorders. Next, all inpatients evaluated by an in-hospital neurologist were screened twice daily for study inclusion and data abstraction was performed by neurology attendings, residents and fellows. COVID-19 patients who were prospectively excluded due to “no new neurological disorder” after evaluation by a neurologist were eligible for inclusion in the control group.

### Neurological Diagnoses and Severity of Illness Scales

Neurological diagnoses --including toxic-metabolic encephalopathy, hypoxic-ischemic encephalopathy, stroke (ischemic or hemorrhagic), seizure, neuropathy, myopathy, movement disorder, encephalitis/meningitis, myelopathy, myelitis—followed established criteria ^11-20^ and were coded for COVID-19 patients found to have a *new* neurological complication (excluding recrudescence or worsening of old neurological deficits) as diagnosed by in-hospital neurology teams. A second review of neurological diagnoses was performed by relevant subspecialty co-authors (e.g. stroke, neurocritical care, epilepsy sub-specialists). Patients could be coded for more than one neurological complication. A control group consisted of a matched cohort of COVID-19 patients *without* new neurological complications admitted during the same time frame (March 10, 2020 and May 20, 2020) and to the same hospitals. Controls were propensity score matched 1:1 to cases by age, gender and intubation status (as a marker of illness severity during index hospitalization) using a matching ratio of 0.01, random order drawing of matches, without replacement and with priority given to exact matches.

Demographic data, past medical history, clinical course and hospital outcomes (mortality rates discharge disposition, ventilator days and hospital length of stay) were collected in both groups. Severity of illness during hospitalization was assessed using both the maximum recorded Sequential Organ Failure Assessment (SOFA) score, and the lung severity scale^21^ for hospitalized patients (0=no oxygen requirement, 1=supplemental oxygen required, 2=high-flow nasal cannula or non-invasive ventilation, 3=invasive mechanical ventilation). Past medical history data was gathered through review of the medical record. A past history of dementia was coded for patients with pre-existing diagnoses of mild cognitive impairment, Alzheimer’s type dementia, vascular dementia, Lewy body/Parkinson’s related dementia, progressive supranuclear palsy, multiple system atrophy, corticobasal degeneration, frontal-temporal dementia, normal pressure hydrocephalus or Creutzfeld-Jakob disease. Pre-morbid baseline mRS were collected based on patient/surrogate report.

### Study Outcomes

Longitudinal 6-month follow-up assessments were conducted by telephone interview among case and control hospital survivors or their surrogates who consented to participate. Contact was attempted at 6-months (±1month)from the onset of neurological symptoms among cases, or from the onset of COVID-19 symptoms among controls. Based on prior data, the median time from general COVID-19 symptom onset to neurological complication was 2 days^10^. Three attempts at contact were required before patients/surrogates were coded as “unreachable”.

We selected outcome measures that could be assessed via telephone interview, were validated for completion by surrogates, and could be completed in approximately 30 minutes to avoid participant fatigue. The primary outcome was the modified Rankin Scale [mRS; 0=no symptoms, 6=dead]^22^, analyzed using an ordinal proportional odds model. Secondary outcomes included: the Barthel Index^23^ for activities of daily living (0=completely dependent, 100=independent for all activities), the Telephone Montreal Cognitive Assessment (MoCA; 22=perfect score; ≤18=abnormal cognition)^24^, and Quality of Life in Neurological Disorders^25^ (Neuro-QoL) short form self-reported health measures of anxiety, depression, fatigue and sleep. Patients with fewer than 13 years of education received an additional point when scoring the telephone MOCA^26^. The outcomes of occurrence of return to work among those employed pre-morbidly and hospital readmission were self-reported by the patient or their surrogate. All of the above batteries have been validated for surrogate completion with the exception of the telephone MoCA, which was only scored if the patient was able to complete the assessment. Incomplete or partial responses to a given metric were excluded from analysis.

### Standard Protocol Approvals and Patient Consents

This study was approved by the NYU Grossman School of Medicine Institutional Review Board. All patients or their surrogates provided consent for participation.

### Statistical Analyses

Demographic variables, past medical history, clinical course and in-hospital outcomes were compared between COVID-19 patients with and without a new neurological event using the Mann-Whitney U test for non-normally distributed continuous variables and Chi-square test for categorical values, as appropriate. Neuro-QoL raw scores were converted into T-scores with a mean of 50 and standard deviation of 10 in a reference population (U.S. general population or clinical sample)^27^. Higher T-scores indicate worse self-reported health for the anxiety, depression, fatigue and sleep metrics. Secondary outcomes were dichotomized based on clinically relevant or published thresholds. For patients who died, a mRS score of 6 was assigned, but no other outcome variables were scored.

Ordinal logistic regression models predicting 6-month mRS were constructed to determine the effect of neurological complications during COVID-19 hospitalization. Secondary outcomes were compared between those with or without neurological disorders using Mann-Whitney U (Wilcoxon rank-sum) or Chi-squared tests as appropriate. Variables (demographics, past medical history, clinical course) predictive of mRS scores with a P value ≤0.100 were entered into multivariable ordinal logistic regression analysis to estimate the adjusted odds ratios (aOR) and 95% confidence intervals (CI) for the primary outcome of mRS score. Clinically relevant interactions were tested. Additionally, multivariable, backward step-wise, binary logistic regression models were constructed predicting dichotomized secondary outcomes of Barthel Index (fully independent versus any abnormal ADLs), MoCA (abnormal score≤18 versus >18^24^), Neuro-QoL T-scores worse than average (>50) and return to work, including univariate predictors with a P-value ≤0.100. Correlations between different outcome measures were assessed using Spearman correlation coefficients. All analyses were conducted using IBM SPSS Statistics for Mac version 26 (IBM Corp., Armonk, NY).

## RESULTS

Of 606 hospitalized COVID-19 patients prospectively identified with new neurological complications, 395 (65%) survived to hospital discharge^10^. Among the 3,885 COVID-19 control patients *without* neurological complications hospitalized during the same time frame, 3,134 (81%) survived and 395 were propensity-score matched by age, gender and ventilator status to the surviving neurological patients (Supplemental Table 1). Seven-hundred-ninety follow-up calls were attempted at 6-months, and 382 (N=196 neurological cases and N=186 controls) were completed (Figure 1). The median time from first COVID-19 symptom (e.g. fever, cough, nausea, vomiting, diarrhea) to neurologic complication onset was 2 days (interquartile range [IQR] 0-13). The median time from neurological symptom onset (or COVID-19 symptom onset for controls) to follow-up interview was 6.7 (IQR 6.5-6.8) months. Between discharge and 6-months, 40/196 (20%) neurological patients and 35/186 (19%) controls died (P=0.715) in a median of 0.8 (IQR 0.4-2.1) months from symptom onset. Among those able to participate in outcome assessments, direct responses were obtained from 121/156 (78%) neurological patients and 131/151 (87%) controls, while surrogate responses were utilized in 35/156 (22%) neurological patients and 20/151 (13%) controls (P=0.026).

**Figure 1.**
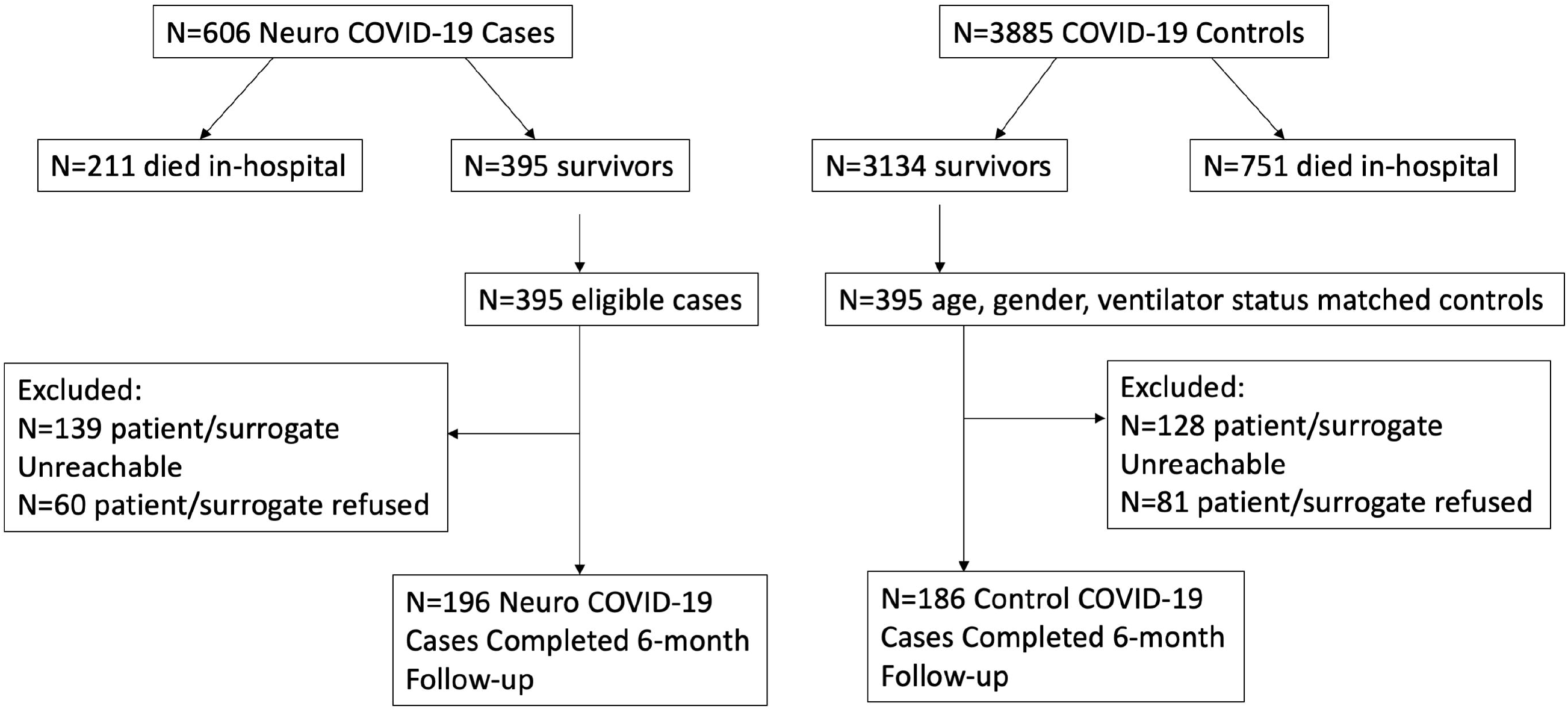
Flow Chart of Case and Control Inclusion and Exclusion

Among the neurological cohort, the most common diagnoses were toxic-metabolic encephalopathy (52%), hypoxic-ischemic encephalopathy (21%), stroke (11%), and seizure (11%, Table 1). Compared to control patients, neurological patients had lower body mass indices, more past history of dementia, stroke and seizure, worse SOFA scores during hospitalization and higher rates of acute renal failure (all P<0.05, Table 1). Patients with newly diagnosed neurological events were more often discharged to a nursing home and less often discharged home than control patients.

**Table 1.** Comparison of COVID-19 patients with and without neurological disorders during hospitalization

| **Characteristic** | **Neurologic**<br>**COVID-19**<br>**N=196** | **COVID-19**<br>**Control**<br>**N=186** |
| --- | --- | --- |
| **Demographics** |  |  |
| Median Age (IQR)-yr | 68 (55-77) | 69 (57-78) |
| Male sex-no./total no. (%) | 128/196 (65%) | 120/186 (65%) |
| Body Mass Index-median (IQR)* | 26 (23-30) | 28 (24-35) |
| Race- no./total no. (%) |  |  |
| White | 86/196 (44%) | 77/186 (41%) |
| Black | 22/196 (11%) | 26/186 (14%) |
| Asian | 20/196 (10%) | 8/186 (4%) |
| Native American/Pacific Islander | 1/196 (0.5%) | 1/186 (1%) |
| American Indian | 1/196 (0.5%) | 1/186 (1%) |
| Other | 25/196 (13%) | 36/186 (19%) |
| Prefer not to answer | 41/196 (21%) | 37/186 (20%) |
| Ethnicity- no./total no. (%) |  |  |
| Hispanic | 30/196 (15%) | 40/186 (22%) |
| Non-Hispanic | 122/196 (62%) | 110/186 (59%) |
| Prefer not to answer | 44/196 (23%) | 36/186 (19%) |
| Past Medical History- no./total no. (%) |  |  |
| Dementia* | 28/196 (14%) | 11/186 (6%) |
| Psychiatric Illness | 32/196 (16%) | 16/186 (9%) |
| Stroke (ischemic or hemorrhagic)* | 40/196 (20%) | 17/185 (9%) |
| Seizure* | 32/196 (16%) | 7/186 (4%) |
| Movement Disorder | 5/196 (3%) | 1/186 (0.5%) |
| Multiple sclerosis/demyelinating disease | 0 | 3/186 (2%) |
| Chronic kidney disease | 33/196 (17%) | 18/186 (10%) |
| Hypertension | 78/196 (40%) | 87/186 (47%) |
| Diabetes | 62/196 (32%) | 48/186 (26%) |
| Coronary artery disease | 36/196 (18%) | 33/186 (18%) |
| COPD/Asthma | 17/196 (9%) | 21/186 (11%) |
| Atrial Fibrillation | 28/196 (14%) | 17/186 (9%) |
| Baseline modified Rankin Score-median (range) | 0 (0-5) | 0 (0-5) |
| **Hospital Course** |  |  |
| Date of Admission -median (IQR) | 4/3/2020<br>(3/28/2020-4/10/2020) | 4/2/2020<br>(3/25/2020-4/11/2020) |
| Intensive care unit vs. non-ICU unit- no./total no. (%) | 69/196 (35%) | 54/184 (29%) |
| Intubation-- no./total no. (%) | 66/196 (34%) | 64/186 (34%) |
| Worst SOFA score- median (IQR)* | 4 (3-10) | 4 (3-6) |
| Lowest Oxygen saturation (%), median (IQR) | 88% (74-92%) | 88% (78-91%) |
| Lung Injury Severity Scale<sup>21</sup>, median (IQR) | 1 (1-3) | 1 (1-3) |
| Lowest Mean Arterial Pressure (mmHg), median (IQR) | 63 (52-72) | 65 (54-73) |
| Acute renal failure- no./total no. (%)* | 43/196 (22%) | 23/186 (12%) |
| Hypotension requiring vasopressors- no./total no. (%) | 58/196 (30%) | 43/186 (23%) |
| **Neurological Disorders during Hospitalization** |  |  |
| Toxic/Metabolic Encephalopathy- no./total no. (%) | 102 (52%) | -- |
| Stroke (any type) - no./total no. (%) | 21 (11%) | -- |
| Ischemic/TIA | 15 (8%) | -- |
| Intracerebral/Intraventricular hemorrhage | 6 (3%) | -- |
| Spontaneous Subarachnoid hemorrhage | 0 | -- |
| Seizure (clinical or electrographic) - no./total no. (%) | 21 (11%) | -- |
| Hypoxic/ischemic brain injury- no./total no. (%) | 42 (21%) | -- |
| Movement Disorder- no./total no. (%) | 15 (8%) | -- |
| Neuropathy- no./total no. (%) | 15 (8%) | -- |
| Myopathy- no./total no. (%) | 7 (4%) | -- |
| Guillain Barre Syndrome- no./total no. (%) | 3 (1.5%) | -- |
| Encephalitis/meningitis- no./total no. (%) | 0 | -- |
| Myelopathy/Myelitis- no./total no. (%) | 0 | -- |
| **Medications during Hospitalization** |  |  |
| Corticosteroids- no./total no. (%) | 49/196 (25%) | 52/186 (28%) |
| Hydroxychloroquine- no./total no. (%) | 138/196 (70%) | 134/186 (72%) |
| Azithromycin- no./total no. (%) | 121/196 (62%) | 125/196 (67%) |
| Remdesivir- no./total no. (%) | 0 | 1/186 (0.5%) |
| Therapeutic anticoagulation- no./total no. (%) | 75/196 (38%) | 59/186 (32%) |
| Tocilizumab- no./total no. (%) | 1/143 (0.7%) | 0 |

| **Hospital Outcomes** |  |  |
| --- | --- | --- |
| Home- no./total no. (%)* | 97/182 (53%) | 119/178 (67%) |
| Acute rehabilitation facility- no./total no. (%) | 21/190 (11%) | 12/178 (7%) |
| Nursing home - no./total no. (%)* | 57/190 (30%) | 36/178 (20%) |
| LTAC- no./total no. (%) | 6/190 (3%) | 4/178 (2%) |
| Length of Stay- median (IQR) | 10.3 (4.8-32.5) | 8.3 (3.7-19.9) |
| Ventilator Days- median (IQR) | 12.6 (5.5-27.6) | 9.0 (2.9-24.7) |

\*Indicates P<0.05; IQR=interquartile range; COPD=chronic obstructive pulmonary disease; SOFA=sequential organ failure assessment; TIA=transient ischemic attack; LTAC=long term acute care hospital

In the entire cohort, 346/382 (91%) patients had at least one abnormal outcome at 6-months: 170/304 (56%) had limited activities of daily living (Barthel<100), 106/215 (50%) had impaired cognition (telephone MOCA<18), the median mRS was 3 (IQR 1-5), 47% (81/154) of those working pre-morbidly were unable to return to work at 6-months. Of those able to complete the Neuro-QoL batteries, 174/280 (62%) demonstrated worse than average scores compared to reference populations (T-score >50) on at least one metric: 128/280 (46%) scored worse than average on anxiety; 105/278 (38%) on sleep; 98/272 (36%) on fatigue, and 71/279 (25%) on depression.

Six-month mRS scores were significantly worse in patients with neurological complications (median 4, IQR 2-5) compared to controls (median 3, IQR 1-4, unadjusted ordinal logistic regression analysis OR 1.57, 95% CI 1.10-2.24, Wald X^2^(1)=6.096, P=0.014; Figure 2). Patients with neurological complications were more likely to have impaired activities of daily living (53% versus 35% of controls, Chi-squared test P=0.002) and were less likely to return to work (41% versus 64% of controls, Chi-squared test P=0.004; Table 2). Though neurological patients scored worse on the telephone MoCA (median 17 versus 18, Wilcoxon rank-sum test P=0.036), after excluding patients with baseline dementia, there was no difference between groups. Neuro-QoL T-scores for anxiety, depression, fatigue and sleep were also similar between both groups. Hospital readmission occurred in 14% of patients in both groups (Table 2). Overall, persistent dyspnea severe enough to limit normal activity occurred in 35% of patients and did not differ between groups.

**Figure 2.**
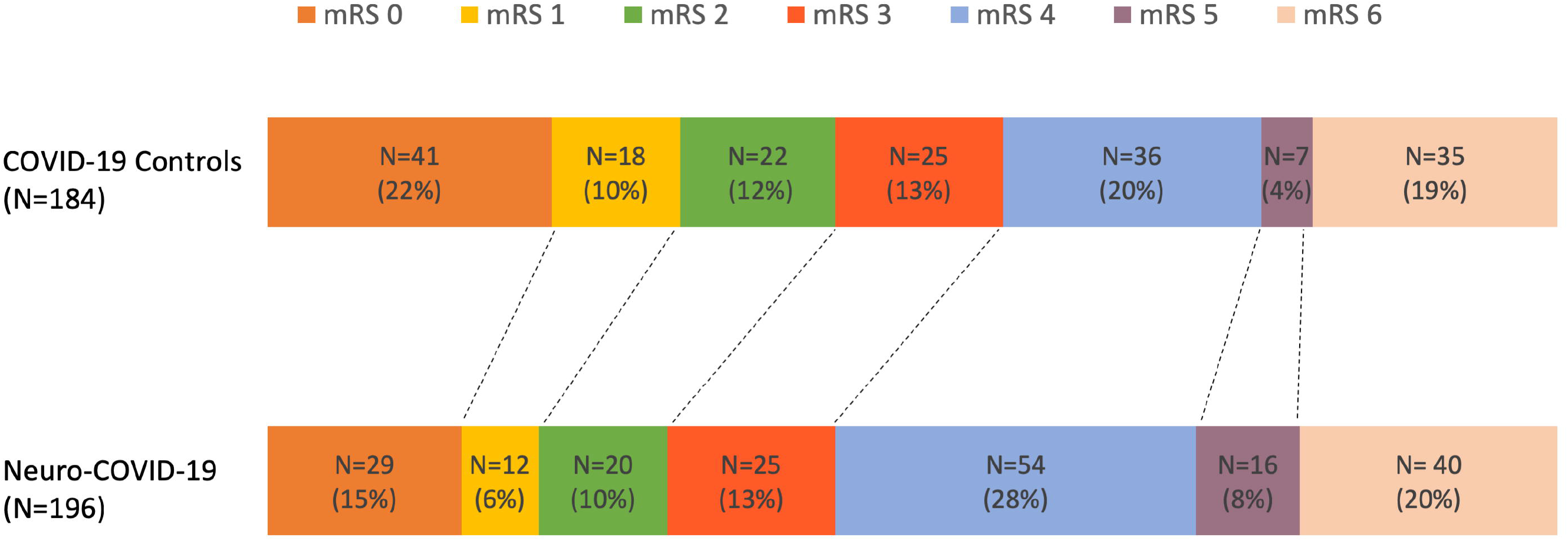
Ordinal Logistic Regression analysis of 6-month modified Rankin scores among patients with and without neurological disorders during hospitalization for COVID-19. (Adjusted odds ratio 2.03, 95% confidence interval 1.22-3.40)

**Table 2.**
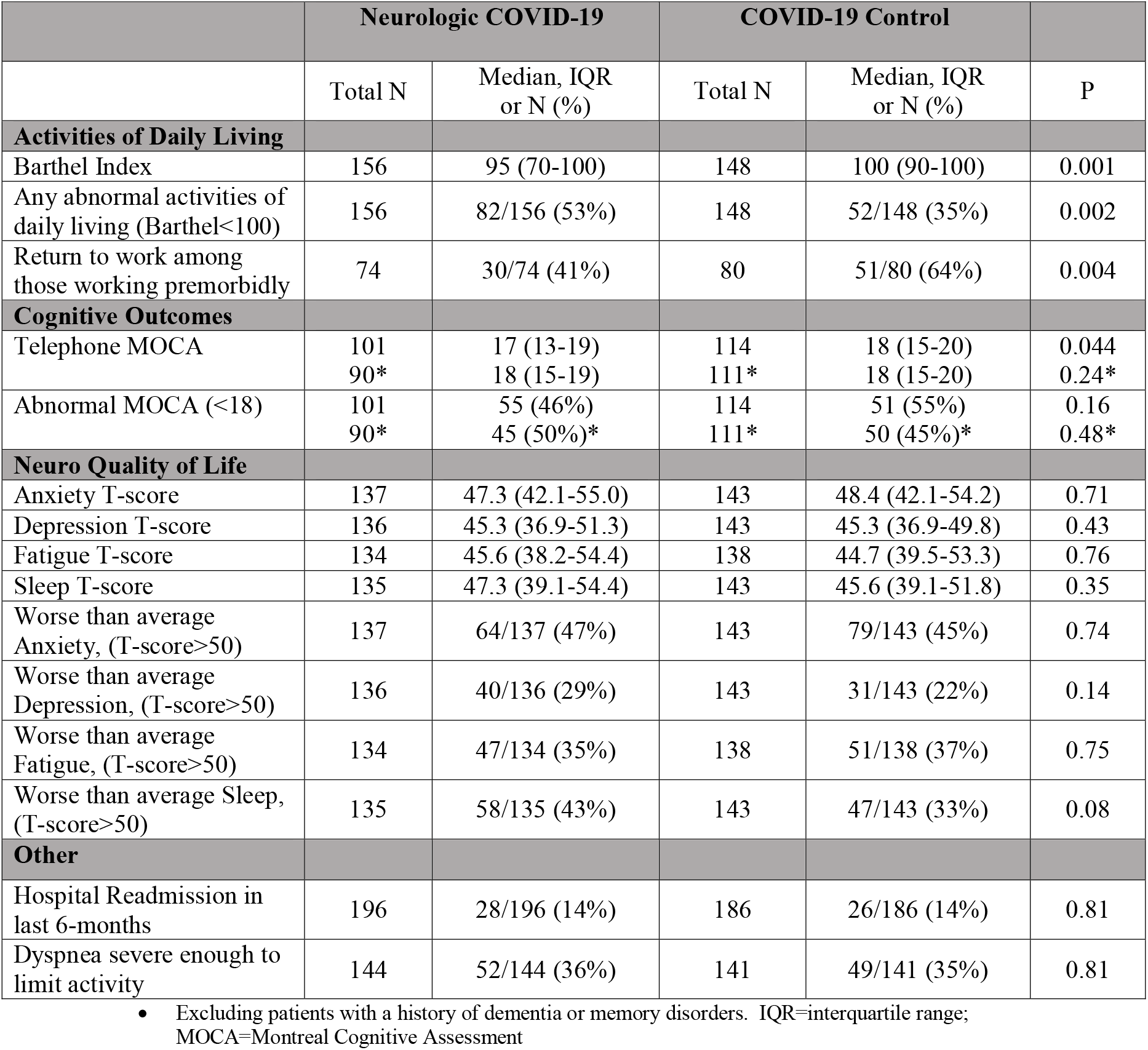
Secondary outcomes at 6-months among COVID-19 patients with and without neurological disorders during hospitalization

|  | Neurologic COVID-19 |  | COVID-19 Control |  |  |
| --- | --- | --- | --- | --- | --- |
|  | Total N | Median, IQR<br>or N (%) | Total N | Median, IQR<br>or N (%) | P |
| **Activities of Daily Living** |  |  |  |  |  |
| Barthel Index | 156 | 95 (70-100) | 148 | 100 (90-100) | 0.001 |
| Any abnormal activities of daily living (Barthel<100) | 156 | 82/156 (53%) | 148 | 52/148 (35%) | 0.002 |
| Return to work among those working premorbidly | 74 | 30/74 (41%) | 80 | 51/80 (64%) | 0.004 |
| **Cognitive Outcomes** |  |  |  |  |  |
| Telephone MOCA | 101<br>90* | 17 (13-19)<br>18 (15-19) | 114<br>111* | 18 (15-20)<br>18 (15-20) | 0.044<br>0.24* |
| Abnormal MOCA (<18) | 101<br>90* | 55 (46%)<br>45 (50%)* | 114<br>111* | 51 (55%)<br>50 (45%)* | 0.16<br>0.48* |
| **Neuro Quality of Life** |  |  |  |  |  |
| Anxiety T-score | 137 | 47.3 (42.1-55.0) | 143 | 48.4 (42.1-54.2) | 0.71 |
| Depression T-score | 136 | 45.3 (36.9-51.3) | 143 | 45.3 (36.9-49.8) | 0.43 |
| Fatigue T-score | 134 | 45.6 (38.2-54.4) | 138 | 44.7 (39.5-53.3) | 0.76 |
| Sleep T-score | 135 | 47.3 (39.1-54.4) | 143 | 45.6 (39.1-51.8) | 0.35 |
| Worse than average Anxiety, (T-score>50) | 137 | 64/137 (47%) | 143 | 79/143 (45%) | 0.74 |
| Worse than average Depression, (T-score>50) | 136 | 40/136 (29%) | 143 | 31/143 (22%) | 0.14 |
| Worse than average Fatigue, (T-score>50) | 134 | 47/134 (35%) | 138 | 51/138 (37%) | 0.75 |
| Worse than average Sleep, (T-score>50) | 135 | 58/135 (43%) | 143 | 47/143 (33%) | 0.08 |
| **Other** |  |  |  |  |  |
| Hospital Readmission in last 6-months | 196 | 28/196 (14%) | 186 | 26/186 (14%) | 0.81 |
| Dyspnea severe enough to limit activity | 144 | 52/144 (36%) | 141 | 49/141 (35%) | 0.81 |

- Excluding patients with a history of dementia or memory disorders. IQR=interquartile range; MOCA=Montreal Cognitive Assessment

While most of the outcome metrics (mRS, Barthel, anxiety, depression, fatigue, sleep, hospital readmission, return to work and persistent dyspnea), with the exception of the telephone MOCA, were significantly correlated with one another, correlation coefficients were generally modest, ranging from Rho=0.155 for readmission and fatigue T-scores, to Rho=0.497 for fatigue and sleep T-scores. However, Barthel and mRS scores were strongly correlated (Rho= −0.788, P<0.001, Supplemental Table 3).

Predictors of 6-month outcomes, including demographics, past medical history, hospital course, specific neurological diagnoses, and COVID-19 specific medications were evaluated in logistic regression analysis (Supplemental Table 2). Results of multivariable ordinal logistic regression analysis for the primary mRS outcome are shown in Table 3 (with all univariate variables in supplemental table 2 with P≤0.100 tested in the model). The occurrence of new neurological complications during COVID-19 hospitalization remained a significant independent predictor of worse 6-month mRS scores (adjusted OR 2.03, 95% CI 1.22-3.40, P=0.01). Other significant predictors were older age, worse baseline functional status (as measured by baseline mRS), and longer hospital length of stay (Table 3). Similarly, neurological complications during hospitalization remained an independent predictor of limited activities of daily living (aOR 0.38, 95% CI 0.20-0.74) and was inversely associated with return to work (aOR 0.31, 95% CI 0.10-0.95; Table 4). Older age, and worse baseline functional status were consistent independent predictors of worse outcome across a spectrum of other outcome measures. Acute respiratory failure requiring invasive mechanical ventilation was associated with limited ADLs and worse than average sleep, while the interaction of hypotension requiring vasopressors and intubation was associated with worse than average fatigue at 6-months, suggesting a synergistic effect of both hypotension and hypoxia (Table 4).

**Table 3.** Predictors of 6-month Modified Rankin Score in multivariable ordinal logistic regression analysis, N=382

| Variable | Adjusted OR (95% CI) | P |
| --- | --- | --- |
| Neurological Complication during Hospitalization | 2.03 (1.22-3.40) | 0.01 |
| Age (per year) | 1.02 (1.01-1.04) | 0.02 |
| Baseline modified Rankin Score | 1.91 (1.58-2.31) | <0.001 |
| Hospital Length of Stay (per day) | 1.04 (1.01-1.06) | 0.01 |

OR=Odds Ratio; CI=confidence interval
Additional Variables tested in the model included: history of atrial fibrillation, history of dementia, history of stroke, history of psychiatric illness, history of chronic kidney disease, history of coronary artery disease, worst sequential organ failure assessment score during hospitalization, toxic metabolic encephalopathy during hospitalization, hypoxic-ischemic encephalopathy during hospitalization, education≤12 years, married versus not, lowest mean arterial blood pressure, use of zinc during hospitalization (all P>0.05)

**Table 4.**
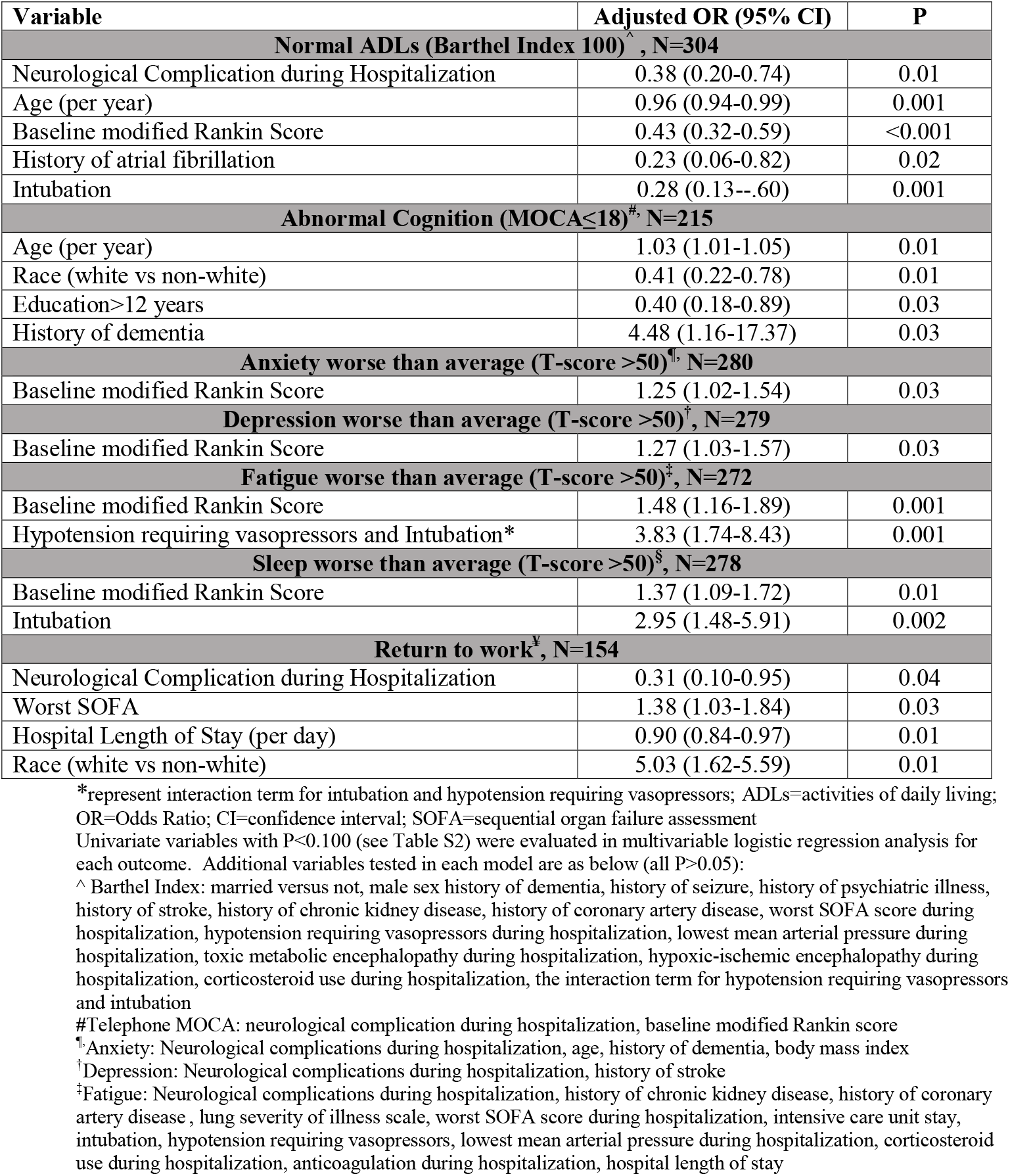
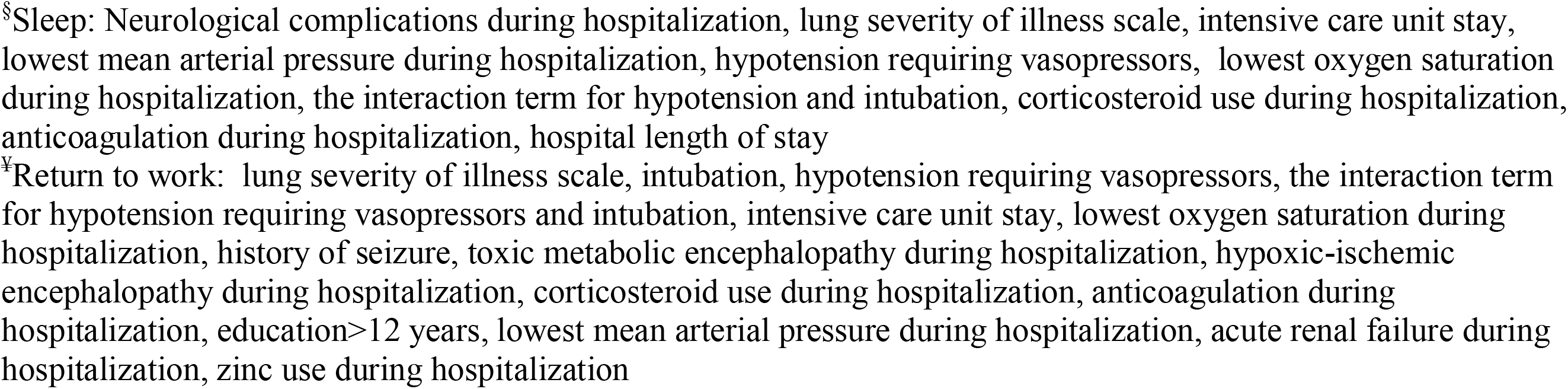
Predictors of 6-month secondary outcomes in multivariable binary logistic regression analysis.

## DISCUSSION

In this prospective study, we found that patients diagnosed with new neurological complications during hospitalization for COVID-19 had a 2-fold increased odds of worse 6-month functional outcome compared to hospitalized COVID-19 patients without neurological complications. Furthermore, over half of these neurologically affected patients could not independently perform some of their basic activities of daily living and 59% of those employed prior to COVID-19 infection were unable to return to work by 6-months. While return to work metrics may be impacted by pandemic-related economic conditions, patients with neurological complications were still significantly less likely to return to work than patients living in the same metropolitan area without neurological complications. While patients with new neurological events had higher baseline levels of dementia, stroke, and seizure, increased odds of worse 6-month functional outcomes persisted after adjusting for these baseline differences, as well as for baseline premorbid modified Rankin scores.

Though we hypothesized that patients with neurological disorders would generally have worse long-term outcomes, we found that both groups of patients had high rates of cognitive impairment approaching or exceeding 50% at 6-months. While we did not compare our patients to non-COVID controls, the estimated prevalence of mild cognitive impairment in the general community among patients aged 65-69 years (the median age of our population) is only 8.4%^28^ and the prevalence of dementia is <1%^29^. Notably, we did not find a correlation between depression and cognition scores, suggesting that depression alone is not the cause of cognitive deficits. Neuropsychiatric symptom burden was also substantial, with nearly two-thirds of all patients reporting worse than average anxiety, depression, fatigue or sleep disorders. Because we used T-scores for Neuro-QoL metrics, these findings represent abnormalities compared to reference populations. Though we identified significant relationships between most outcome measures and self-reported persistent dyspnea, the association was weak and it does not seem that dyspnea alone can fully account for the levels of disability or cognitive impairment that we measured.

Strengths of our study include its prospective ascertainment of data beginning at the time of index hospitalization, initial neurological diagnoses by board-certified neurologists^10^, a propensity-score matched control group and a multi-domain outcome battery. This is the first study, to our knowledge to prospectively assess long-term neurological function and cognitive outcomes among COVID-19 hospital survivors in a standardized fashion. Other studies have presented short-term outcomes between 2 weeks and 2 months from COVID-19 symptom onset, but neither used formal assessments of fatigue, mood, cognition or functional status^30,31^. In one study of outpatients diagnosed with COVID-19, reports of protracted symptoms during the subacute phase (14-21 days) included persistent fatigue in 35%, persistent loss of taste/smell in 20% and confusion in 20%^31^. Another study in a hospitalized COVID-19 cohort found that 87% of patients had at least one lingering COVID-19 symptom in a mean of 60 days after initial COVID-19 symptom onset, with the most common long-term symptoms being fatigue (53%) and dyspnea (43%)^30^. In a large, 6-month study of a COVID-19 cohort from Wuhan, China^21^, fatigue or muscle weakness was reported in 63%, and sleep difficulties in 23%, however, these metrics were not assessed using a standardized battery and it is unclear how these rates compare to a reference population. This study also identified anxiety or depression in 23% based on the EuroQol five-dimension five-level questionnaire, but this metric does not differentiate anxiety and depression. This study also excluded some of the sickest patients such as those with underlying dementia, psychiatric illness, hospital readmission, nursing home or facility patients and those physically unable to return for an in-person follow-up. This may explain the higher rates of anxiety (46% of patients), and depression (25%) that we observed.

We identified neurological complications during index COVID-19 hospitalization, age, and worse premorbid baseline functional status as independent predictors of poor outcomes, however, these risk factors are not readily modifiable. Invasive mechanical ventilation and hypotension requiring vasopressors were associated with worse activities of daily living, fatigue and sleep. These variables may be potentially modifiable by medications and therapeutics that were not available early in the pandemic. While we anticipated an association between the severity of lung injury, hypoxia and worse cognitive outcomes based on prior ARDS studies^32^ and neuropathological findings of hypoxic brain injury in COVD-19 autopsy specimens^33-37^, we did not find a significant relationship. One possibility is that the telephone MOCA may not be sensitive enough to detect certain types of cognitive impairment. We used this tool because it is validated for phone interview and would allow us to gather data on patients that would not otherwise be able to complete follow-up in person due to pandemic related restrictions. Another possibility is that we were unable to fully capture the duration and severity of hypoxemia burden using stochastic measures of the lowest oxygen saturation or the lung injury severity scale^21^.

Furthermore, there is likely an interaction between hypoxemia and hypotension that compounds the risk of cognitive deficits, however, we only identified a significant effect of the interaction of intubation and hypotension requiring vasopressors on the outcome of fatigue. The degree and duration of hypoxemia and/or hypotension required to cause permanent brain injury may vary from patient to patient depending on the presence of flow-limiting extra- or intracranial vessel stenosis, carbon dioxide levels, the integrity of cerebral autoregulation, prior ischemic damage, and the degree of brain metabolic activity and blood flow coupling. Big data studies evaluating oxygen and blood pressure levels are needed to evaluate these associations further.

We also documented a high rate of readmission (14%) in both the neurological patients and controls compared to other studies, which reported rates as low as 1% at 6-months^21^ and 3.6% at 14-days post discharge^38^. However, the first study excluded patients for whom follow-up would be difficult (including those readmitted to a hospital for underlying diseases)^21^ and the other looked at short-term readmissions within its own hospital system, not accounting for readmissions that could occur outside of its network^38^. Because our readmission data came directly from patients or their surrogates, we were able to capture readmissions across different hospital systems. It is likely that limited access to outpatient follow-up during the New York COVID-19 surge may have contributed to this high readmission rate. Systematic video and phone follow-up early after discharge may help mitigate readmission risk.

There are limitations to our work. First, though all patients completed the mRS evaluation, the sickest patients were unable to complete other metrics due to limited function. Thus, secondary outcomes may actually be worse than what we were able to measure. Additionally, family members of patients who did poorly may be less motivated to participate in research. Second, certain outcomes may be confounded by unmeasured factors. For example, return to work may be impacted by the current economic environment. Anxiety, depression, fatigue and sleep may be similarly impacted by unmeasured societal, economic and environmental pandemic-associated factors. Comparing COVID-19 patients to similar community members without COVID-19 would help quantify the impact of some of these unmeasured factors. Third, though we identified high rates of disability among both neurological COVID-19 patients and controls, this study was not designed to determine whether the deficits measured by these metrics are static or represent ongoing injury as suggested by some “long-hauler” and post-COVID literature^39^. Since cognitive impairment and mood abnormalities occur in over 50% of ARDS survivors^40^, it is possible that the level of disability we are documenting is not specific to SARS-CoV-2, but rather a result of factors related to critical viral illness. Additional one-year follow-up in this cohort may help clarify the trajectory of recovery. Fourth, the telephone MOCA is meant to screen for cognitive impairment, but more extensive neuropsychological testing is needed to identify specific domains of cognitive abnormality. While we did adjust for years of education when scoring the telephone MOCA, other societal factors may impact scores^26^. Finally, while we did find a small, yet significant relationship between ongoing severe dyspnea and other outcome metrics, we did not use a standardized measure of dyspnea because we felt that a longer survey would lead to participant fatigue and incomplete responses. Further investigation of the impact of long-term pulmonary sequelae on neurological outcomes is warranted.

## CONCLUSIONS

Over 90% of COVID-19 patients who survived hospitalization had at least one abnormal outcome metric at 6-months: 56% of patients had limited activities of daily living, 50% had abnormal cognition, 62% had worse than average anxiety, depression, fatigue or sleep and 47% of those working pre-morbidly were unable to return to work. Patients with neurological complications during their index COVID-19 admission had significantly worse functional outcomes as measured by the mRS and Barthel scales, and were less likely to return to work, even after accounting for other premorbid factors. The frequency and severity of functional, cognitive and mood abnormalities among COVID-19 hospital survivors--with or without neurological complications--should factor into educational and preventive efforts directed at patients, physicians and the general public.

## Data Availability

De-identified data will be made available to qualified investigators upon written request to the corresponding author.

## Acknowledgments

We thank the patients and families who agreed to participate in this study.

